# Critical timing for triggering public health interventions to prevent COVID-19 resurgence: a mathematical modelling study

**DOI:** 10.1101/2021.07.06.21260055

**Authors:** Zhuoru Zou, Christopher K Fairley, Mingwang Shen, Nick Scott, Xianglong Xu, Zengbing Li, Rui Li, Guihua Zhuang, Lei Zhang

## Abstract

To prevent the catastrophic health and economic consequences from COVID-19 epidemics, some nations have aimed for no community transmission outside of quarantine. To achieve this, governments have had to respond rapidly to outbreaks with public health interventions. But the exact characteristics of an outbreak that trigger these measures differ and are poorly defined. We used existing data from epidemics in Australia to establish a practical model to assist stakeholders in making decisions about the optimal timing and extent of interventions. We found that the number of reported cases on the day that interventions commenced strongly predicted the size of the outbreaks. We quantified how effective interventions were at containing outbreaks in relation to the number of cases at the time the interventions commenced. We also found that containing epidemics from novel variants that had higher transmissibility would require more stringent interventions that commenced earlier. In contrast, increasing vaccination coverage would enable more relaxed interventions. Our model highlights the importance of early and decisive action in the early phase of an outbreak if governments aimed for zero community transmission, although new variants and vaccination coverage may change this.

## Introduction

The coronavirus disease 2019 (COVID-19) pandemic continues to cause catastrophic health and economic crisis around the world^1,2^. To prevent the consequences of the COVID-19 epidemic, 18 vaccine candidates have been approved by the World Health Organization^3^. Yet, achieving global herd immunity with these vaccines will take time, and existing vaccines also face challenges from emerging SAR-CoV-2 variants with mutations that evade vaccine-elicited immune response^4^. Non-pharmaceutical interventions remain the most effective means for COVID-19 control until herd immunity can be achieved. Non-pharmaceutical interventions have been successful in countries such as New Zealand, Australia, China, and Singapore. These countries all share one thing in common; that early intervention results in more effective control of outbreaks.

Outbreak surveillance for early COVID-19 detection is essential to inform control measures. It enables stakeholders to commence interventions in time to avoid lengthy and extensive restrictions down the track and minimise health and economic losses. Several studies have explored surveillance indicators for the early detection of COVID-19 outbreaks, including COVID-19-related digital data streams and SARS-CoV-2 viral fragments detection in wastewater^5,6^. Modelling studies have also informed stakeholders by projecting the trends and severity of COVID-19 epidemics in the context of different policy decisions^7–11^. However, none of these studies quantified the commencement time and extent of interventions when facing a COVID-19 outbreak of unknown severity and impact to the community. A predictive model that provides timely alerts for intervention commencement would be of great practical value in curbing a COVID-19 outbreak.

Our objective was to establish a predictive model to assist stakeholders in decision-making regarding timely intervention commencement based on limited surveillance data in the early stages of an outbreak. This model integrates existing public health interventions (including contact tracing, social distancing, face mask use, voluntary testing, and vaccination). It predicts the severity of the COVID-19 epidemic in the near future depending on the extent and timing of interventions. This allows outbreaks to be contained. We selected Australia as a case study because it is one of the few countries that dramatically limit the health and economic consequences of the COVID-19 epidemic through a zero community transmission approach. Our study finding would provide an early outbreak warning to intervention commencement in Australia and is transferrable to other settings worldwide.

## Results

### Historical outbreaks of the COVID-19 epidemic and R_e_

We identified nine local outbreaks in the four selected Australian states from 25 January 2020, when the first COVID-19 case was reported in Australia, to 12 March 2021 (Table 1, Fig. 1). We observed that the reported cases on the day interventions commenced were highly predictive of the subsequent size of the epidemic peak and duration of the outbreak (Fig. 2, Spearman correlation, ρ1=0.97, ρ2=0.78, both p<0.05). Of the nine outbreaks, four times the state government intervened when the number of daily reported cases was below 10 cases, and the subsequent peak size of the outbreak was limited (1–10 cases), and the outbreak was contained within a month. In contrast, on four occasions, the state governments intervened when the number of daily cases was between 10–30 cases. The subsequent outbreak peak was substantial (<100 cases), and the outbreak was contained within three months. On one occasion, intervening late at a daily reported case of 149 resulted in a very high outbreak peak of 687 cases and an outbreak duration of almost five months (Table 1, Fig. 1–2).

**Table 1.**
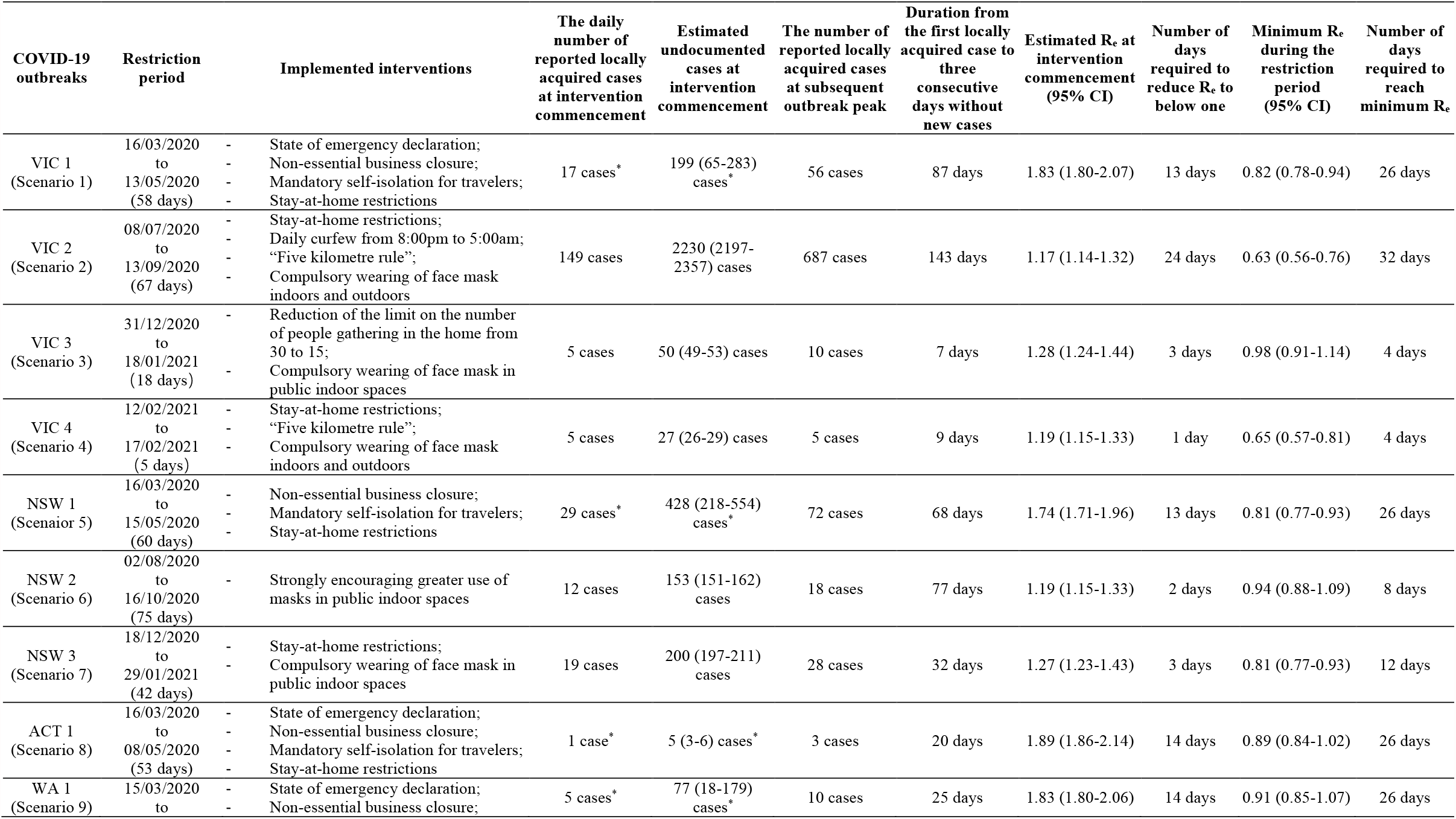
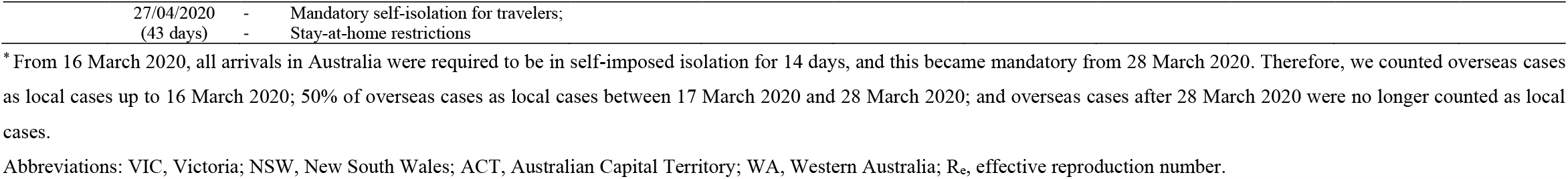
Nine COVID-19 outbreaks in the four Australian states.

**Fig. 1.**
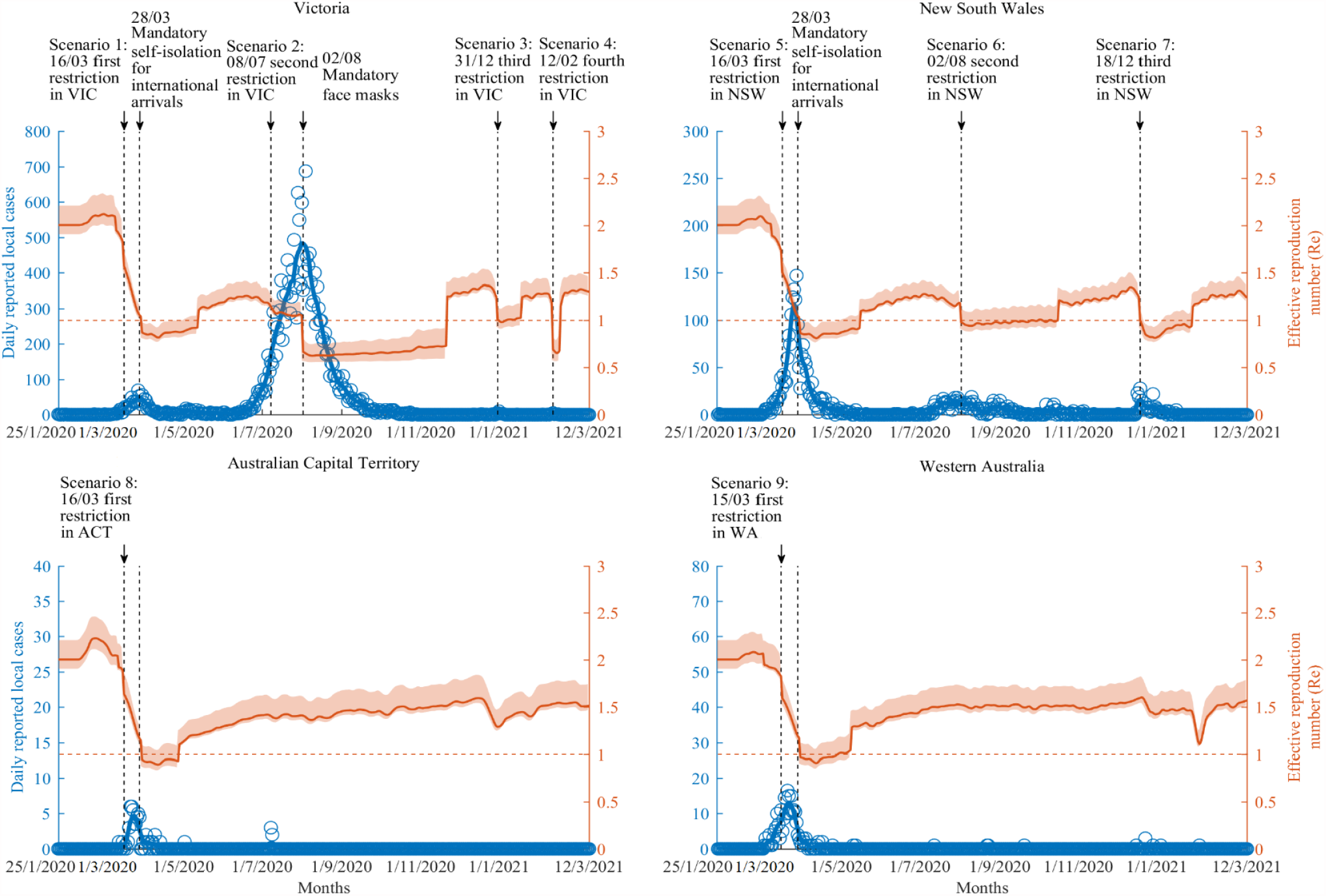
Historical outbreaks of the COVID-19 epidemic and R_e_ in four Australian states (25 January 2020 – 12 March 2021)

**Fig. 2.**
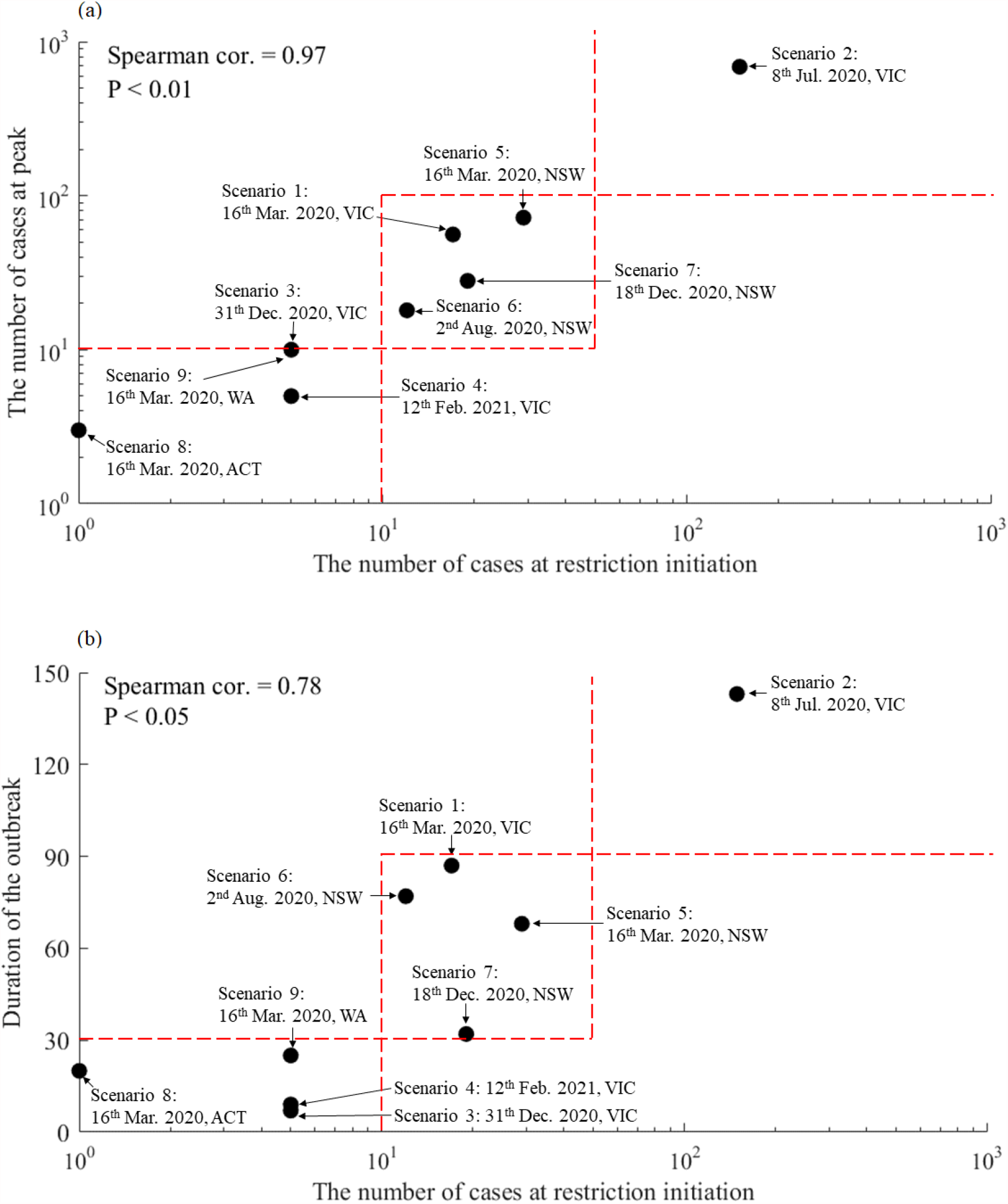
**Association between the reported cases on the day interventions commenced and the subsequent peak size and duration of the outbreak**

At a baseline scenario with contact tracing, voluntary testing, but no vaccination, we estimated the effective reproduction number (R_e_) of COVID-19 to be 1.88 (95% CI: 1.85–2.13) in Australia. We observed that the interval between intervention commencement and reduction of R_e_ to below one was shortened with increasing experience in responding to the COVID-19 epidemic (Table 1, Fig. 1). In four recent outbreaks (after the second outbreak in Victoria), R_e_ was reduced to near or below one less than one week after the intervention commencement.

### Surveillance for outbreak severity

Fig. 3a illustrated the prediction of the potential outbreak severity based on current daily reported cases and public health interventions. The first panel illustrated how the combinations of various levels of reduction in social activity and face mask coverage might impact on R_e_ under the baseline scenario. We observed that in the Australian setting, reducing social activity by two-third of the pre-epidemic level or increasing face mask use to at least 77% would reduce R_e_ to below the threshold curve of one. The second panel demonstrated the projected average number of daily cases over the next 7 days based on various combinations of R_e_ and the actual numbers of daily reported cases. It showed three distinct regions that reflect various epidemic severities. Region A, where R_e_≥1, indicated an uncontrolled and expanding epidemic in the near future; Region B, where R_e_ < 1 but the predicted average daily cases over the next 7 days would still exceed a manageable level (e.g. 10 cases/day), indicated a controlled epidemic with a substantial risk of resurgence; Region C, where R_e_ < 1 and the predicted average daily cases over the next 7 days were <10 cases, indicated a controlled epidemic with a reducing risk of resurgence.

**Fig. 3.**
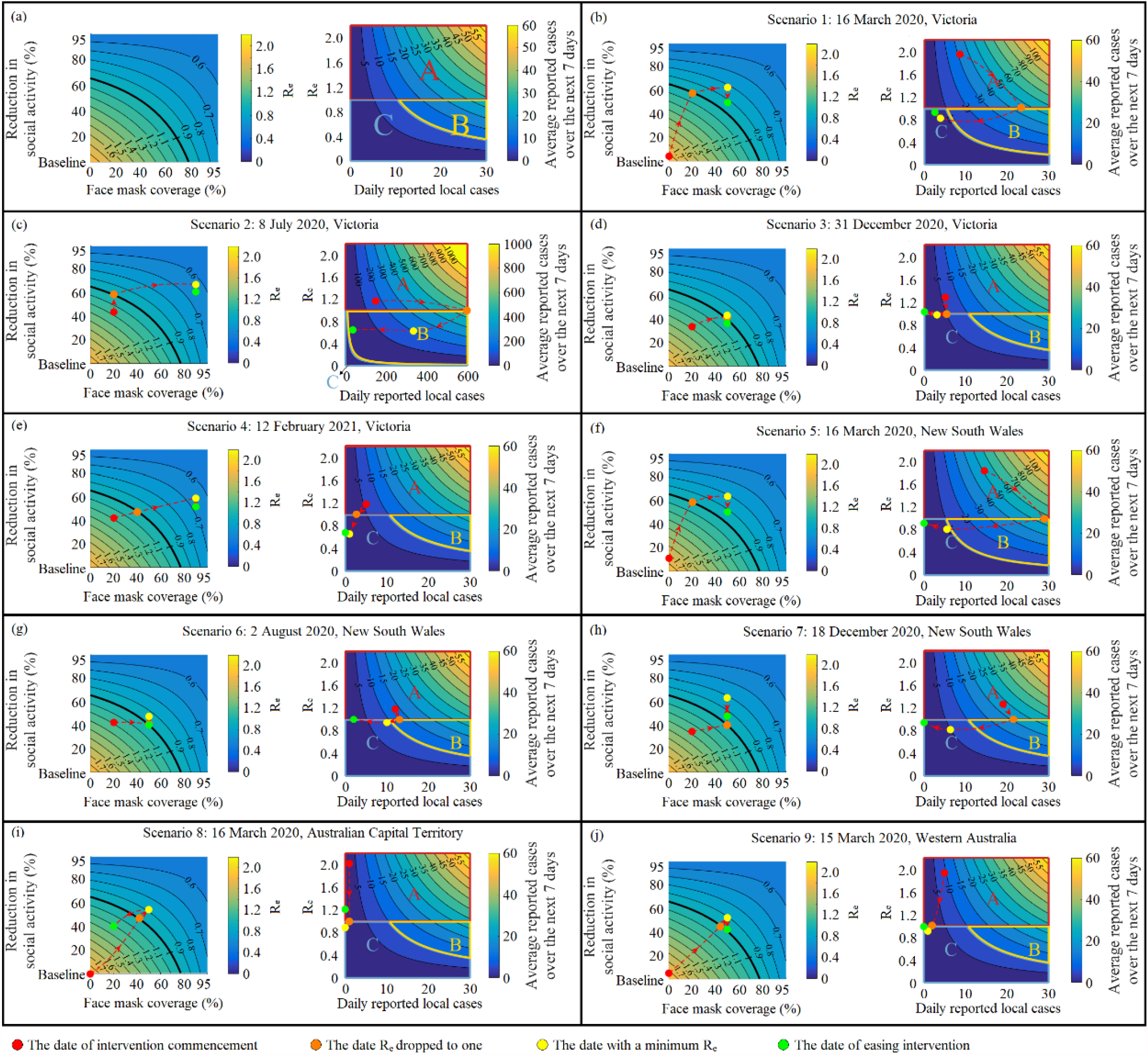
**Trajectories of changes of intervention, R_e_ and the predicted outbreak severity for the past COVID-19 outbreaks in four Australian states**

Fig. 3b–j illustrated the trajectories of intervention change and the predicted outbreak severity for the above nine outbreaks, from the date of intervention commencement to the date R_e_ dropped to one, to the date with a minimum R_e_, and to the date of easing intervention. We observed that on five occasions, the trajectory of the projected epidemic severity underwent a shift from region A to region B before moving to region C, suggesting a substantial risk of a larger and longer-lasting outbreak. In contrast, on the remaining four occasions, the trajectory of the projected epidemic severity shifted directly from region A to region C, suggesting a well-controlled outbreak. These were consistent with the actual outbreak outcomes (Table 1, Fig. 1–2). More importantly, the trajectories of intervention change did not differ substantially across the nine occasions (all reduced R_e_ below one), and the number of cases on the day intervention commenced largely determined whether the trajectory of epidemic severity would pass through region B.

### Critical timing for intervention commencement to avoid major outbreaks

We predicted the average number of daily cases over the first 7 days after interventions at various combinations of the number of daily reported cases and R_e_ on the day of intervention commencement (Fig. 4). We imposed a condition that the interventions of social distancing and face mask use would reduce R_e_ uniformly to one within 7 days. We found that in the baseline scenario, to contain the epidemic to an average of ≤10 daily cases over the next 7 days, the critical number of daily reported cases that should trigger interventions would be 6 cases.

**Fig. 4.**
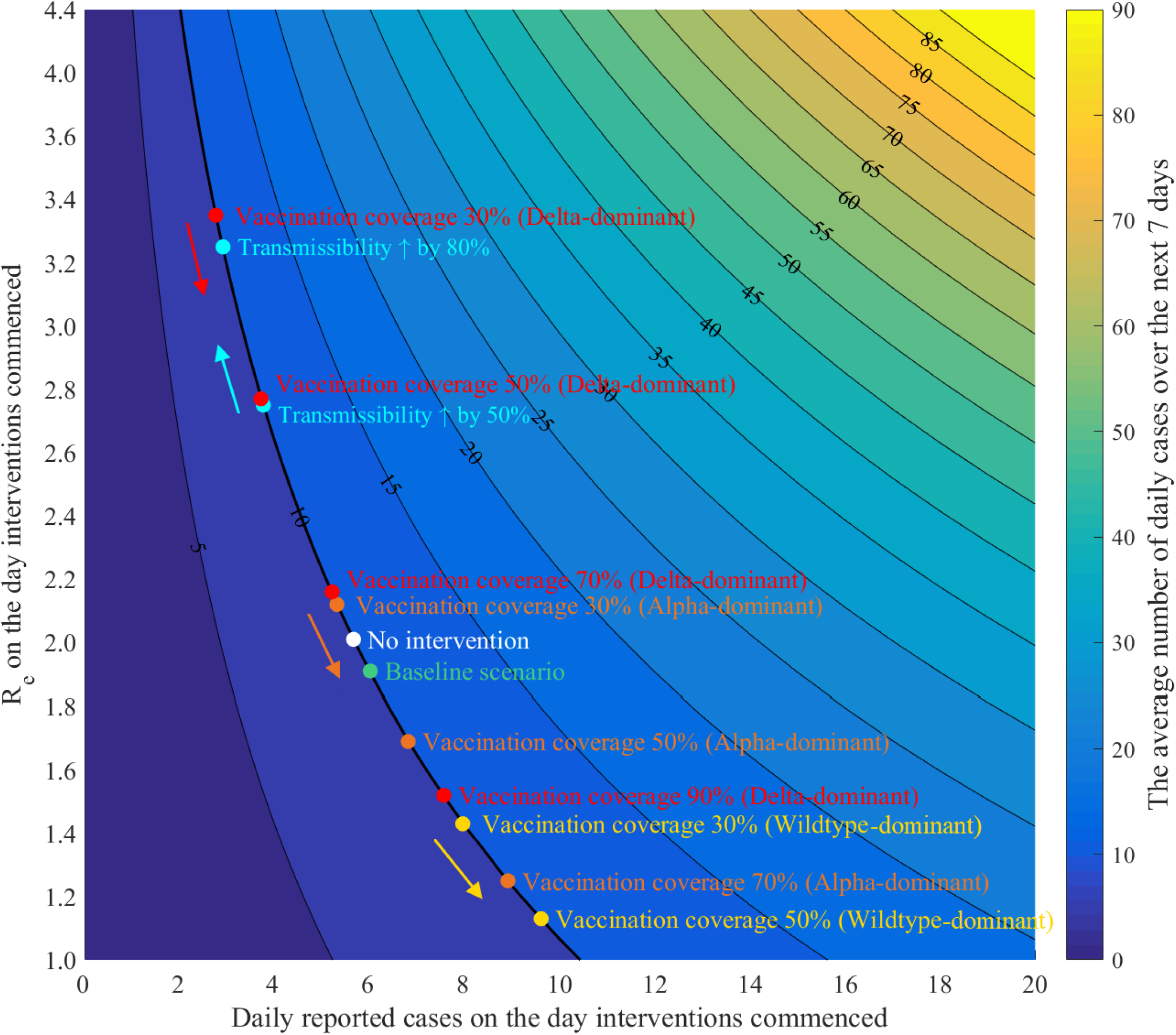
**Predicted outbreak severity at various combinations of the number of daily reported cases and R_e_ on the day of intervention commencement**

### Impact of viral variants on R_e_ and critical timing for intervention commencement

If the transmissibility of the novel variants increased by 50%, 80%, 110%, and 140%, R_e_ would be increased to 2.75, 3.25, 3.73, and 4.20, respectively. Consistently, we observed a rising or even vanishing threshold curve of R_e_ (R_e_=1, Fig. 5). It indicated that more stringent combinations of social distancing and face mask use, even in combinations with vaccination, would be required to reduce R_e_ below one. Further, the critical timing for intervention commencement would need to be advanced as the transmissibility of the variants increases (Fig. 4). With a 50% (estimated 40–80% for the Alpha variant^12–14^) increase in transmissibility and no vaccination, reducing social activity by 80% of the pre-epidemic level combined with mandatory masks (50% coverage) could reduce R_e_ to one. The critical number of cases to trigger intervention commencement would be limited to 4 cases to maintain the average number of daily cases over the next 7 days to ≤10 cases (Fig. 4–5). In contrast, with a 140% (estimated to be 60% higher for the Delta variant than for the Alpha variant^15^) increase in transmissibility, social distancing and face mask use alone would not be sufficient to reduce R_e_ below one.

**Fig. 5.**
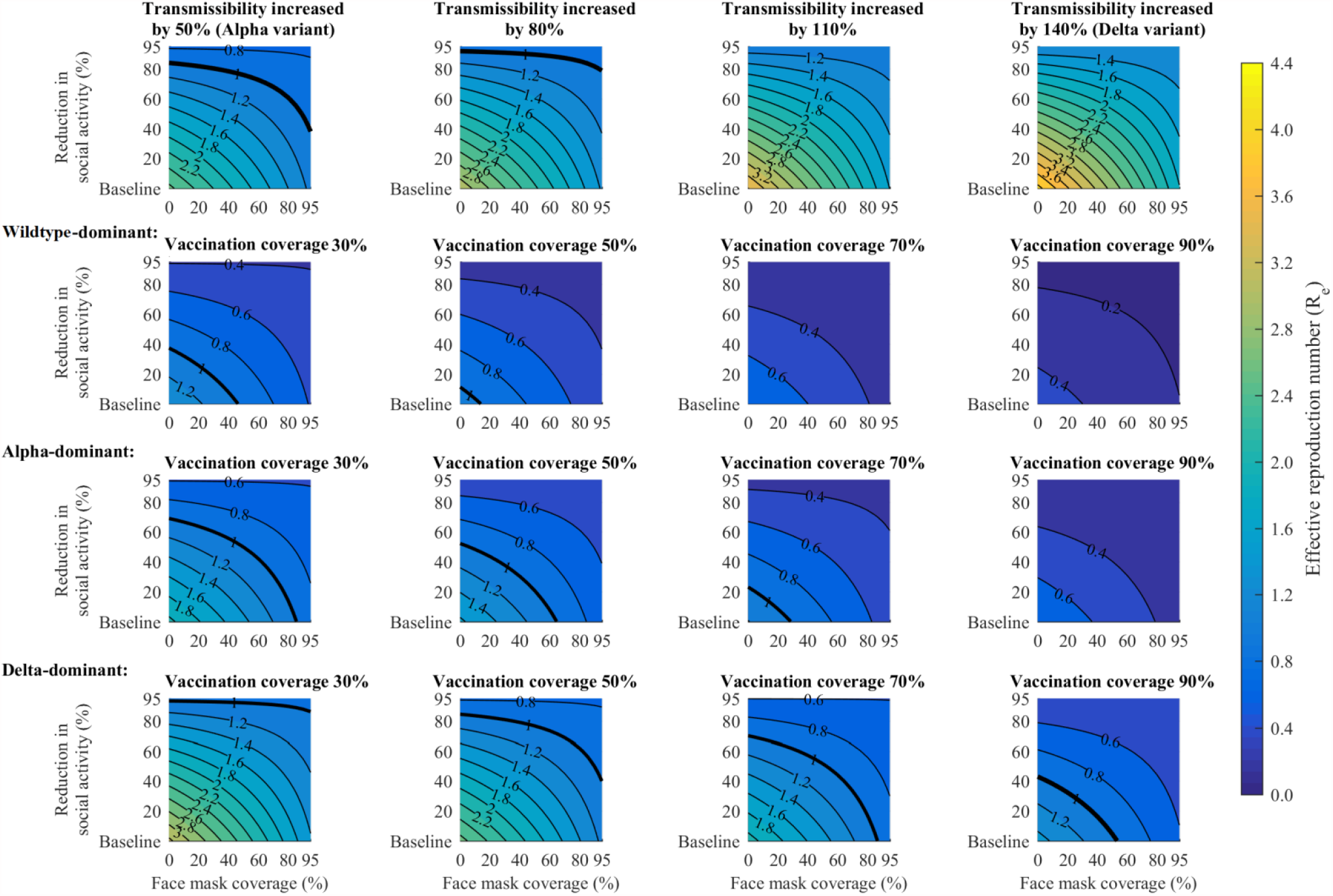
Effect of various levels of reduction in social activity and face mask coverage on Re under different scenarios.

### Impact of vaccination on R_e_ and critical timing for intervention commencement

If COVID-19 vaccination coverage reach 30%, 50%, 70% and 90% in a wildtype dominant epidemic, R_e_ would be significantly reduced to 1.43, 1.13, 0.82, and 0.51, respectively (Fig. 5). We also observed a substantial decrease in the threshold curve of R_e_ (R_e_=1). The curve would disappear if vaccination coverage exceeds 70% (Fig. 5), indicating that social distancing and face mask use restrictions would not be necessary if 70% of Australians are vaccinated. However, in an Alpha-dominant epidemic, vaccination coverage would need to reach 90% for social distancing and mask use restrictions to be fully relaxed. In a Delta-dominant epidemic, combinations of vaccination with social distancing and face mask use restrictions were required to reduce R_e_ to one. Further, in all wildtype and variants dominant epidemics, the critical timing for intervention commencement could be delayed with increasing vaccination coverage (Fig. 4).

## Discussion

Our study identified the critical timing and extent for commencing public health interventions to contain COVID-19 outbreaks in an Australian setting. We found that in the past outbreaks in four Australian states, the number of reported cases on the day interventions commenced was a strong predictor of the subsequent peak size of the outbreaks. We quantified the effective combinations of interventions that would reduce R_e_ below one and the critical number of daily cases for the commencement of interventions. We indicated that in the early phase of an outbreak, containing the prospective epidemic to a low level (≤10 cases/day) required effective interventions to be commenced before the number of daily reported cases reaches 6 cases. However, an increase in viral transmissibility would require stricter interventions to be commenced with a lower number of daily cases. In contrast, increasing vaccination coverage would allow for less strict interventions and an increase in the threshold for their commencement.

Our study developed a practical model to assist decisions for determining the optimal timing and extent of interventions from a stakeholders’ perspective. To our best knowledge, this study is the first of its kind to integrate existing public health interventions and epidemic severity to quantify the risk of COVID-19 resurgence. It provides intuitive recommendations to stakeholders to take early and decisive action. Our study also quantified the impact of various potential changes in viral transmissibility and levels of vaccination on the timing and extent of interventions for future adjustments, which is an important consideration given the current epidemic of the Delta variant worldwide. Additionally, the model may be extended to demonstrate future epidemic trends resulting from various combinations of different levels of public health interventions commenced at different time points and may be transferable to other countries facing resurgent COVID-19 epidemics by adjusting appropriate parameters.

Our study confirmed that the early commencement of strong public health interventions is critical in containing a COVID-19 outbreak. Our findings are echoed in many outbreaks in other settings. For example, in two outbreaks in New Zealand in August 2020 and February 2021, the government immediately declared an elevated alert level with only 4 and 3 cases per day, respectively, resulting in rapid containment of both outbreaks and a return to no community transmision^16,17^. Similar interventions were initiated very early and aggressively in both Vietnam and Taiwan, and both regions showed success in their control of the first wave of the epidemic^18^.

Our results are particularly relevant when facing the emergence of more transmissible SARS-CoV-2 variants. The Alpha variant reported in the UK in late 2020 has about 50% (range: 40–80%) higher transmissibility than the wildtype^12–14^. It has become dominant in the United States and many parts of Europe, leading to a rebound of the epidemic^19,20^. For this variant, our model predicts that stricter measures that include 80% reduction in social activities and 50% public face mask use would be necessary to contain the epidemic without vaccination, and these restrictions need to be implemented earlier when there are only 4 reported daily cases. The recent report of the Delta variant demonstrated an even higher (60%) transmissibility than that of the Alpha variant^15^. That implies that combinations of vaccination and other interventions would be necessary to contain it (Fig. 5). However, our study encouragingly illustrated that expanded COVID-19 vaccination is an effective way for COVID-19 control and socioeconomic recovery in the future. Nevertheless, concerted efforts of other public health interventions are still necessary if high coverage of vaccination cannot be guaranteed in the short term or facing more transmissible new variants.

Our study has several limitations. First, we simulated historical epidemics based on four Australian states but excluding Queensland, which experienced a significant outbreak in March–April 2020. That is because the state’s official reports did not include information on whether a diagnosed case is from a known or unknown source and hence cannot inform our model. Second, the rate of viral transmission may vary across countries due to differences in health care capacities and the extent of public health interventions. Our study uses Australia as a case study but intends to generalise this work to other settings in future investigations. Similarly, we did not consider environmental differences, which is likely to play a role and differ between states. Third, to the completion date of this study, reliable data on the transmissibility and mortality of the novel variants of SARS-CoV-2 and the effectiveness of the COVID-19 vaccine against new variants are still under investigation. Our study was conducted with limited availability of these data. Finally, we did not differentiate between the nature of the new cases. Cases in the same household as a known case and those who were isolated at the time of their diagnosis will be different from cases that are not linked to known cases and who were not isolated at the time of their diagnosis. It was not possible to incorporate these qualitative differences in a quantitative model, but they clearly matter.

In conclusion, our study quantified, for the first time, the extent of public health interventions that would effectively control an outbreak and the critical timing for the commencement of interventions. It provides stakeholders with intuitive recommendations to assist them in taking early and decisive action and is, therefore, has important implications for facilitating the achievement of the ambitious goal of rapid and complete control of the COVID-19 outbreak.

## Methods

### Data source

We collected COVID-19 epidemic data, including the number of daily reported cases (both with known and unknown sources), cumulative confirmed cases, and deaths in Victoria, Australia (25 January 2020 – 12 March 2021) based on official reports from the Victorian Department of Health and Human Services. We calibrated our model against the Victorian data. We also searched and collected similar data (if available) from other Australian states, including New South Wales, Australian Capital Territory, and Western Australia, for model validation. We collected relevant health policies and intervention implementation timelines in response to the COVID-19 epidemic from the official website of the Australian Government Department of Health by states and social activity data from Google COVID-19 community mobility data^21^ (Supplementary Fig. 2–4).

### Model structure and assumptions

We constructed a Susceptible-Infected-Recovered compartmental model to simulate the transmission of SARS-CoV-2 in the Australian population. The model consisted of ten compartments (Supplementary Fig. 1). Susceptible individuals could be infected through contact with an infectious source in public places and households. The force of infection was dependent on the infectivity of the virus, the presence and severity of symptoms of the infectious source, the venue of exposure, and public health interventions in the population (Supplementary Information p4–5). The model assumed that approximately 17% of the infected individuals were asymptomatic and never developed any noticeable symptoms^22^. Asymptomatic individuals would not be isolated but later spontaneously recovered unless they were diagnosed due to voluntary testing or contact tracing. The remaining 83% infected individuals would be ‘pre-symptomatically infected’ with an incubation period (5.8 days on average)^23^, then later become symptomatic. Symptomatic individuals would be diagnosed and isolated after an interval from symptom onset to isolation (4.2 days on average)^24^. We assumed diagnosed individuals would die of COVID-19-related complications with a certain probability. Transition probabilities in the model were shown in Supplementary Table 1.

### Interventions in the model

We simulated five public health interventions in the model, including contact tracing, social distancing, face mask use, voluntary testing, and vaccination.

Contact tracing enabled back-tracing of the individuals who were in close contact with diagnosed individuals. We simulated the effect of contact tracing in three steps. First, we estimated the number of close contacts for newly diagnosed individuals in public places and households based on the definition of close contacts^25,26^. Second, the number of infected individuals among close contacts was estimated according to the force of infection. According to the model, infected close contacts would enter the asymptomatic/pre-symptomatic infection compartments. Uninfected close contacts would enter the susceptible compartment. Third, depending on the effectiveness of contact tracing (i.e., the ability to detect and quarantine all close contacts), a proportion of infected close contacts would be diagnosed and isolated, while a proportion of uninfected close contacts would be tested and quarantined but later returned to the susceptible compartment. The effectiveness of contact tracing may depend on various factors, such as willingness to cooperate, recall bias, availability of contact tracers, and quarantine compliance. Previous reports have indicated that 20% of the close contacts would be uncooperative, and among those who cooperate, 60% of recollected information may be incorrect^27,28^. Also, a further 20% of close contacts would likely fail to comply with quarantine^27^. Considering the above unavoidable factors, we estimated that approximately 80% of close contacts among the remaining close contacts (cooperative and without recall bias) were detected and isolated in Australia through model calibration (Supplementary Information p5–6, 10).

Voluntary testing was given to individuals who believed they were in close contact with infected individuals and may be at risk of infection. According to the cumulative number of COVID-19 voluntary tests over the past 7 days and the population size, we estimated that 0.09%–0.2% of the Australian population would receive voluntary testing each day. We hence assumed the same proportion of asymptomatic/pre-symptomatic individuals would voluntarily test each day and be subsequently diagnosed and isolated (Supplementary Information p6, 11).

Social distancing would reduce the average number of daily contacts in the public space. Based on proportional deviations of real-time mobility from the pre-epidemic level in public places from ‘Google COVID-19 community mobility data’, we estimated the average number of contacts at various levels of social distancing restrictions. Further, we estimated the daily probability of exposure to infectious individuals in public places (Supplementary Information p4–5, 9–10).

Face mask use would reduce the probability of transmission in each exposure, which would be reduced by 85% (95% CI: 50–95%) in a single exposure with the presence of a face mask^29,30^ (Supplementary Information p4–5, 10).

Vaccination would reduce the proportion of susceptible individuals in the population. According to Australia’s vaccine agreements, the Pfizer/BioNTech vaccine (40 million doses available), the Oxford/AstraZeneca vaccine (53.8 million doses available), and the Moderna vaccine (25 million doses available) will account for 33.7%, 45.3%, and 21.0% of COVID-19 vaccination in Australia, respectively^31^. The efficacy of the Pfizer/BioNTech vaccine, the Oxford/AstraZeneca vaccine, and the Moderna vaccine has been reported to be 95% (90.3–97.6%), 67.1% (52.3–77.3%), and 94.1% (89.3–96.8%), respectively^32–34^. We hence estimated the weighted population vaccination effectiveness to be about 82.2%. Recent studies have shown that the existing vaccines remain equally effective in preventing clinical severities in patients infected with Alpha and Delta variants. Still, there was a slight decrease in effectiveness against infection. The efficacy of the Pfizer/BioNTech vaccine reduced to 93.4% (90.4–95.5%) for Alpha and 87.9% (78.2–93.2%) for Delta variant^35–37^. The efficacy of the Oxford/AstraZeneca vaccine reduced to 66.1% (54.0–75.0%) for Alpha and 59.8% (28.9–77.3%) for Delta variant^35–37^. We, therefore, assumed a 2% and 10% reduction in the efficacy of the vaccine against Alpha and Delta (Supplementary Information p6, 11).

### Model simulation and calibration

We calibrated the model by comparing the model-simulated epidemic indicators with the observed data, including the number of daily reported cases, the number of daily known-source cases, the number of daily unknown-source cases, and cumulative deaths in Victoria. We employed a ‘genetic algorithm’ to search within the model parameters’ initial ranges to determine the ‘optimal’ parameter set based on the goodness-of-fit score and the likelihood ratio test^38,39^. We performed about 1,000,000 simulations in total. We ranked the goodness-of-fit scores of the ‘good-fitting sets’ in ascending order and retained the top 1000 sets of best goodness-of-fit. The comparison of the model outputs of the top 1000 parameter sets with the corresponding observed data were shown in Supplementary Fig. 5. The best-fitting set was introduced into the model as the base-case values of model parameters (Supplementary Table 1), and the top 1000 parameter sets were used for sensitivity analysis. All analyses and simulations were performed in MATLAB R 2019a (Supplementary Information p11–13).

### Implications of daily reported cases with known and unknown sources

When a COVID-19 case was diagnosed, contact-tracers would adopt a backwards-tracing strategy to identify the source of transmission. A diagnosed case could be linked to the source of infection if (1) the source was symptomatic and had been diagnosed, or (2) the source was pre-symptomatic/asymptomatic but had been diagnosed through voluntary testing, or (3) the source was pre-symptomatic and was diagnosed due to the onset of symptoms during contact tracing. A diagnosed individual could have an unknown source of infection only when the source was asymptomatic and had not been detected by voluntary testing. With these assumptions, the actual reported number of unknown-source cases might reflect the number of potentially asymptomatic infections. In contrast, the number of known-source cases might reflect the number of pre-symptomatic/symptomatic infections in the population. The reasoning progress was graphically depicted in Supplementary Fig. 7.

To quantify the above relationships, we explored the association between reported daily locally acquired cases and model-estimated potential undocumented infections using data from the second outbreak in Victoria (1 June 2020 – 30 October 2020). Data from this period were selected for analysis because (1) since all international passenger flights were cancelled in Victoria during the second outbreak, this largely excluded the impact of cases from overseas, and (2) this was the most severe of the COVID-19 outbreaks in Australia and data were adequate. An approximately linear relationship was found between the number of daily unknown-source cases and the number of estimated active asymptomatic infections with the Pearson correlation coefficient of 0.741 (linear regression, b = 13.32 [IQR: 12.9–14.14], p < 0.001). A significant linear relationship was found between the number of daily known-source cases and the number of estimated active pre-symptomatic/symptomatic infections with a Pearson correlation coefficient of 0.92 (linear regression, b = 13.13 [IQR: 12.96–13.9], p<0.001) (Supplementary Fig. 8–9).

### Key model outputs

#### Undocumented infections during outbreaks

Undocumented cases represented a potential risk of further community transmission of SARS-CoV-2. Three types of infections were considered ‘undocumented’ in our model. They were asymptomatic infections, pre-symptomatic infections, and symptomatic infections before diagnosis (Supplementary Fig. 6).

#### Basic and effective reproduction number

The basic reproduction number (R_0_) quantified the transmissibility of an infectious pathogen in its initial phase. It was defined as the average number of secondary cases generated by a single infectious case introduced into a fully susceptible population^40,41^. Our model calculated the Next-Generation Matrix^42^ to derive expressions of R_0_ and estimated R_0_ to be 2.01 (1.91–2.21) in Australia, consistent with the previous finding of 1.40–2.27^43–45^. The effective reproduction number (R_e_) measured changes in the actual transmissibility of an infectious disease in a population with the implementation of public health interventions^46,47^. We presented the time-varying R_e_ of COVID-19 by the Australian state from 25 January 2020 to 12 March 2021 in Fig. 1. More detailed derivation and calculation of R_0_ and R_e_ were presented in the Supplementary Information p13.

#### Predicted number of reported cases in the next 7 days

We assessed the risk of the COVID-19 outbreak by predicting the number of secondary cases potentially caused by undocumented cases over the next 7 days. In this setting, we assumed all documented cases were well isolated and would not lead to further community transmission. We estimated the number of secondary infections caused by the currently undocumented cases as follows. First, we estimated R_e_ under the influence of the current public health interventions (Fig. 1), representing the average number of secondary cases caused by an undocumented case during an average infectious period of 14 days. The 14 days was estimated by weighting the average interval from infection to isolation for symptomatic individuals and the average interval from infection to spontaneous recovery for asymptomatic individuals (Supplementary Information p16). Second, the number of undocumented cases was estimated based on the daily number of unknown-source and known-source cases through the above linear relationships. Third, the overall number of secondary cases over the next 14 days was obtained by multiplying R_e_ with the number of undocumented cases. Based on data from past outbreaks, we found that dividing this number by 2 gave a good fit for the number of reported cases over the next 7 days (Supplementary Information p16–17).

#### Critical timing for intervention commencement

We used the number of daily reported locally acquired cases as an indicator to inform the critical timing for intervention commencement. Previous studies had reported that the turning point of the epidemic would occur after a delay of approximately one week from the intervention commencement^48–50^. At the same time, the number of daily reported cases would likely continue to rise until R_e_ was effectively reduced below one. The turning point marked the peak of the outbreak. Therefore, for the time gap between intervention commencement and when R_e_ fell below one, it would be in the best interest for the stakeholders to confine the number of daily reported cases to a manageable level (e.g. ≤10 cases/day) to avoid overloading its care capacity or substantially impact on country’s economic activities. We defined the number of reported cases that would trigger intervention commencement to reduce R_e_ to below one and maintain the average number of daily reported cases over a 7-day period into interventions to be the critical timing for intervention implementation.

We assumed that R_e_ would decrease uniformly from its initial value to one after 7 days into the interventions. We used an autoregressive approach to estimate the average number of reported cases 7 days into the intervention. First, we projected the average daily number of cases over the next 7 days using the number of reported locally acquired cases and R_e_ on the day of intervention commencement. We assumed that the average daily number of cases equals the number of cases on the 4^th^ day; we then linearly interpolated the cases to obtain the number of reported cases one day after intervention. We used a similar approach iteratively to estimate the number of reported cases over the first 7 days after interventions. The critical timing was obtained by limiting the estimated average daily number of cases over the next 7 days after interventions to an average of ≤10 cases/day.

### Model Validation

To validate the reliability and generalisability of the model predictions, we compared the projected number of cases over the next 7 days with the actual number of reported cases over a 7-day period in each of Victoria, New South Wales, Australian Capital Territory, and Western Australia, for the period 25 January 2020 –12 March 2021(Supplementary Fig. 10). The predictions matched the actual epidemic trends well, with an R-squared of 0.99, 0.85, 0.74, and 0.87, respectively.

### Uncertainty and sensitivity analysis

Probabilistic sensitivity analysis was conducted based on 1000 simulations to accommodate the uncertainty of model parameters and determine the 95% confidence interval of the reproduction number. In addition, multiple scenarios were established to explore the impact of the effectiveness of transmissibility of SARS-CoV-2 variants and vaccination coverage on R_e_, the predicted number of reported cases over the next 7 days, and the critical timing of intervention commencement.

## Data Availability

The data that supports the findings of this study are available in the supplementary material of this article.

## Author contributions

L. Z. and Z. Z. designed the study. Z. Z. designed and constructed the model. L. Z., M. S., and N. S. contributed to provide technical and modelling advice throughout the project. Z. Z. performed the modelled analyses, graphed, and interpreted the results. Z. Z., X. X., Z. L., and R. L. contributed to the collection of data and model parameters. Z. Z. drafted the manuscript. L.Z., C.K.F, and G.Z. critically revised the manuscript. All authors reviewed the manuscript and approved the final version.

