## Supplementary Materials for "Critical timing for triggering public health interventions to prevent COVID-19 resurgence: a mathematical modelling study"

### 1. Model equations and detailed description

#### 1.1 Model Structure

We constructed a dynamic compartmental model to describe the spread of SARS-CoV-2 and the impact of public health interventions. The population is divided into ten compartments (Supplementary Fig. 1): susceptible individuals (S), asymptomatic infections (A), pre-symptomatic infections (E), symptomatic infections before diagnosis (I), diagnosed individuals with isolation and treatment (T), uninfected individuals among the quarantined close contacts ($Q_S$), infected individuals among the quarantined close contacts ($Q_{EA}$), vaccinated individuals (V), recovered individuals (R), and dead individuals (D). The total population size is denoted by N, where $\mathrm{N} = \mathrm{S} + \mathrm{A} + \mathrm{E} + \mathrm{I} + \mathrm{T} + Q_S + Q_{EA} + V + R$.

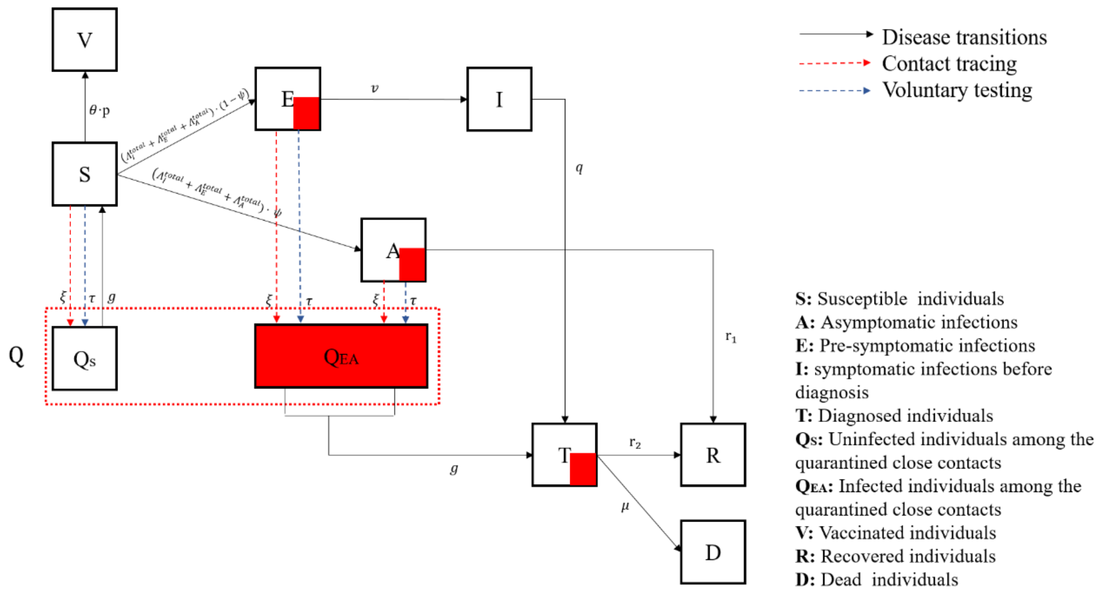


**Supplementary Fig. 1 A schematic flow diagram of the transmission of SARS-CoV-2**

#### 1.2 Model equations

The model is described by the following system of ordinary differential equations. The symbols are defined in the legend following the equations.

$$
\begin{aligned}
\dot{S} &= -\Lambda_I^{total} - \Lambda_E^{total} - \Lambda_A^{total} - Cont_{non-infection} \cdot \xi - \tau \cdot S - \theta \cdot p \cdot S + g \cdot Qs \\
\dot{A} &= \left(\Lambda_I^{total} + \Lambda_E^{total} + \Lambda_A^{total}\right) \cdot \psi - r_1 \cdot A - Cont_{infection} \cdot \psi \cdot \xi - \tau \cdot A \\
\dot{E} &= \left(\Lambda_I^{total} + \Lambda_E^{total} + \Lambda_A^{total}\right) \cdot (1-\psi) - v \cdot E - Cont_{infection} \cdot (1-\psi) \cdot \xi - \tau \cdot E \\
\dot{I} &= v \cdot E - q \cdot I \\
\dot{Q_S} &= Cont_{non-infection} \cdot \xi + \tau \cdot S - g \cdot Q_S \\
\dot{Q_{EA}} &= Cont_{infection} \cdot \xi + \tau \cdot (E + A) - g \cdot Q_{EA} \\
\dot{T} &= q \cdot I + g \cdot Q_{EA} - r_2 \cdot T - \mu \cdot T \\
\dot{R} &= r_1 \cdot A + r_2 \cdot T \\
\dot{D} &= \mu \cdot T
\end{aligned}
\qquad (1)
$$

Parameters in the differential equations:

| Compartments symbols | Description |
|---|---|
| $S$ | Susceptible individuals |
| $A$ | Asymptomatic infected individuals (cases who never developed any noticeable symptoms during the entire period of their disease) |
| $E$ | Pre-symptomatic infected individuals (cases who have mild symptoms before the onset of symptoms) |
| $I$ | Symptomatic but undiagnosed individuals |
| $Q_S$ | Uninfected individuals among the quarantined close contacts |
| $Q_{EA}$ | Infected individuals among the quarantined close contacts |
| $T$ | Diagnosed individuals with isolation and treatment |
| $R$ | Recovered individuals |
| $D$ | Individuals who died from COVID-19-related complications |
| **Parameter symbols** | **Description** |
| $\Lambda_I^{total}$ | Probability of being infected by exposure to symptomatic infected individuals (I) in public places and households |
| $\Lambda_E^{total}$ | Probability of being infected by exposure to pre-symptomatic infected individuals (E) in public places and households |
| $\Lambda_A^{total}$ | Probability of being infected by exposure to asymptomatic infected individuals (A) in public places and households |
| $\psi$ | The proportion of asymptomatic infections among newly infected individuals |
| $1/v$ | The mean incubation time (days) |
| $1/q$ | The interval from symptom onset to isolation in hospital or quarantine (days) |
| $1/g$ | The interval from testing to diagnosis (days) |
| $1/r_1$ | The mean time from infection to recovery for asymptomatic infected individuals (days) |
| $1/r_2$ | The mean time from diagnosis to recovery for symptomatic infected individuals (days) |
| $\mu$ | Disease-induced death rate |
| $Cont_{non-infection}$ | Uninfected close contacts of the daily new confirmed cases |
| $Cont_{infection}$ | Infected close contacts of the daily new confirmed cases |
| $\xi$ | Effectiveness of contact tracing |
| $\tau$ | The coverage rate of voluntary testing |
| $\theta$ | Efficacy of the COVID-19 vaccine |
| $p$ | The coverage rate of COVID-19 vaccination |

##### 1.3 Modelling disease progression

Newly infected individuals would enter the asymptomatic infection compartment (A) and the pre-symptomatic infection compartment (E) according to the proportions $\psi$ ($0 \leqslant \psi \leqslant 1$) and ($1-\psi$), respectively. In the absence of any public health intervention, asymptomatic infected individuals (A) were assumed to recover naturally at the rate $r_1$. Individuals in the incubation period (E) would progress to the symptomatic infection compartment (I) at the rate $v$. Symptomatic infected individuals (I) were assumed to be diagnosed at the rate $q$ and enter the treatment compartment (T) and then be isolated and treated. We also assumed strict isolation so that isolated individuals could not further infect others. Treated individuals would recover at the rate $r_2$ or die due to the disease at the rate $\mu$.

##### 1.4 Force of infection, social distancing, and face mask use

Susceptible individuals may be infected through contact with undocumented cases (sources of infection), including asymptomatic infected individuals (A), pre-symptomatic infected individuals (E), and symptomatic but undiagnosed individuals (I). Force of infection ($\Lambda^{total}$) is given by the sum of probabilities of being infected by exposure to undocumented cases. That is,

$$\Lambda^{total} = \Lambda_A^{total} + \Lambda_E^{total} + \Lambda_I^{total} \tag{2}$$

Transmission may occur in public places and households. Thus, the probability of being infected by exposure to asymptomatic infected individuals ($\Lambda_A^{total}$), for example, is the sum of probabilities from these two routes. It can be expressed as:

$$\begin{aligned}
\Lambda_A^{total} &= \Lambda_A^{fam} + \Lambda_A^{pub} = \beta_{EA}^{fam} \cdot \frac{A}{N_f} \cdot S + \beta_{EA}^{pub} \cdot c_p(t) \cdot \frac{A}{N} \cdot S \\
\Lambda_E^{total} &= \Lambda_E^{fam} + \Lambda_E^{pub} = \beta_{EA}^{fam} \cdot \frac{E}{N_f} \cdot S + \beta_{EA}^{pub} \cdot c_p(t) \cdot \frac{E}{N} \cdot S \\
\Lambda_I^{total} &= \Lambda_I^{fam} + \Lambda_I^{pub} = \beta_I^{fam} \cdot \frac{I}{N_f} \cdot S + \beta_I^{pub} \cdot c_p(t) \cdot \frac{I}{N} \cdot S
\end{aligned} \tag{3}$$

Where,

$$\begin{aligned}
\beta_I^{fam} &= \beta, & \beta_I^{pub} &= \beta \cdot (1-\rho) \cdot (1 - \sigma \cdot m(t)) \\
\beta_{EA}^{fam} &= \beta_I^{fam} \cdot (1-\varepsilon), & \beta_{EA}^{pub} &= \beta_I^{pub} \cdot (1-\varepsilon)
\end{aligned} \tag{4}$$

Using the example of being infected by asymptomatic infected individuals ($\Lambda_A^{total}$), for household exposure, the probability of being infected ($\Lambda_A^{fam}$) is equal to the risk of coming from a household with an asymptomatic infected individual ($\frac{A}{N_f}$) multiplied by the average daily probability of being infected in the household ($\beta_A^{fam}$); for public places exposure, the probability of being infected ($\Lambda_A^{pub}$) is equal to the probability of being exposed to a person who is an asymptomatic infected individual ($\frac{A}{N}$) multiplied by the average daily probability of being infected by contact with an asymptomatic infected individual in public places ($\beta_A^{pub}$), and multiplied by the average number of contacts in public places per day ($c_p(t)$). Social distancing restrictions will affect $c_p(t)$ and thus prevent infection.

Here $N_f$ denotes the total number of households, which is equal to the total population size (N) divided by the average household size ($c_f$) in Australia. We assumed that undocumented cases are dispersed among different households. Because the prevalence of COVID-19 in Australia is low and the probability of two or more household members being infected simultaneously in various public venues is small. We abbreviated the average daily probability of being infected by contact with the symptomatic infected individual in the household ($\beta_I^{fam}$) as $\beta$. Usually, the average frequency of daily person-to-person contacts in public places is less than that within the home. We, therefore, assumed that the average daily probability of being infected by contact with a symptomatic infected individual in a public place ($\beta_I^{pub}$) is less than that of being infected by contact with a symptomatic infected individual at home ($\beta_I^{fam}$), and donated the percentage reduction as $\rho$ ($0 \leqslant \rho \leqslant 1$). The parameters $\sigma$ and $m(t)$ denote the effectiveness and coverage rate of face mask use in public places, respectively, which reflect the effect of face mask use on infection prevention. We assumed that for contacts with asymptomatic/pre-symptomatic infected individuals, the average probability of being infected is lower, i.e. $(1-\varepsilon)\cdot\beta$ where $0 \leqslant \varepsilon \leqslant 1$ denotes the reduction in daily transmission probability.

##### 1.5 Contact tracing

The number of close contacts of newly diagnosed cases in public places and households was calculated based on the following formula:

$$
\begin{aligned}
Cont_I^{fam} &= I \cdot q \cdot \left(c_f - 1\right) \\
Cont_I^{pub} &= I \cdot q \cdot c_p(t) \cdot t_1 \\
Cont_{EA}^{fam} &= Q_{EA} \cdot g \cdot \left(c_f - 1\right) \\
Cont_{EA}^{pub} &= Q_{EA} \cdot g \cdot c_p(t) \cdot t_2
\end{aligned}
\tag{5}
$$

Where $Cont_I^{fam}$ and $Cont_I^{pub}$ indicate the total number of close contacts in households and public places for new cases diagnosed due to symptoms. Similarly, $Cont_{EA}^{fam}$ and $Cont_{EA}^{pub}$ indicate the total number of close contacts in households and public places for new asymptomatic/pre-symptomatic cases diagnosed due to testing and quarantine. The total number of close contacts was estimated by multiplying the number of new diagnoses by the average number of close contacts per individual. $I \cdot q$ and $Q_{EA} \cdot g$ denote the number of daily new diagnosed cases detected from symptomatic infected individuals and asymptomatic/pre-symptomatic infected individuals, respectively. For each confirmed case, the number of close contacts from the household is the number of family members other than the case, i.e., $c_f - 1$; the number of close contacts from public places is the product of the average daily number of close contacts in public places ($c_p(t)$) and the number of tracing days. The parameters $t_1$ and $t_2$ represent the number of tracing days for symptomatic cases and asymptomatic/pre-symptomatic cases. According to the CDC[1], "an infected person can spread SARS-CoV-2 starting from 2 days before they have any symptoms (or, for asymptomatic patients, two days before the positive specimen collection date) until they meet the criteria for discontinuing home isolation". Thus, we assumed that $t_1$ is equal to the interval from symptom onset to isolation in the hospital ($1/q$) plus two days; $t_2$ is equal to two days.

We estimated the number of infected close contacts ($Cont_{infection}$) and uninfected close contacts ($Cont_{non-infection}$) according to the force of infection, expressed as:

$$
Cont_{infection} = \beta_I^{fam} \cdot Cont_I^{fam} + \beta_I^{pub} \cdot \frac{1}{t_1} \cdot Cont_I^{pub} + \beta_{EA}^{fam} \cdot Cont_{EA}^{fam} + \beta_{EA}^{pub} \cdot \frac{1}{t_2} \cdot Cont_{EA}^{pub} \tag{6}
$$

$$Cont_{non-infection} = \left(1-\beta_I^{fam}\right)\cdot Cont_I^{fam} + \left(1-\beta_I^{pub}\right)\cdot\frac{1}{t_1}\cdot Cont_I^{pub} + \left(1-\beta_{EA}^{fam}\right)\cdot Cont_{EA}^{fam} + (1-\beta_{EA}^{pub})\cdot\frac{1}{t_2}\cdot Cont_{EA}^{pub}$$

($\beta_I^{fam}\cdot Cont_I^{fam} + \beta_{EA}^{fam}\cdot Cont_{EA}^{fam}$) denote the average daily number of infected close contacts in households and ($\beta_I^{pub}\cdot\frac{1}{t_1}\cdot Cont_I^{pub} + \beta_{EA}^{pub}\cdot\frac{1}{t_2}\cdot Cont_{EA}^{pub}$) denote the average daily number of infected close contacts in public places.

Depending on the effectiveness of contact tracing (i.e., the ability to detect and quarantine all close contacts, denoted as $\xi$), a proportion of infected close contacts ($Cont_{infection}\cdot\xi$) would be diagnosed and isolated, while a proportion of uninfected close contacts ($Cont_{non-infection}\cdot\xi$) would be tested and quarantined but later returned to the susceptible compartment. The parameter $\xi$ may depend on various factors, such as willingness to cooperate, recall bias, availability of contact tracers, and quarantine compliance. Previous reports have indicated that 20% of the close contacts would be uncooperative, and among those who cooperate, 60% of recollected information may be incorrect[2,3]. Also, a further 20% of close contacts would likely fail to comply with quarantine[2]. Considering the above unavoidable factors, we estimated that the proportion of remaining close contacts who could be detected and isolated through the capability of contact tracing would be about 80% in Australia through model calibration.

##### 1.6 Voluntary testing and vaccination

Voluntary testing was given to individuals who believed they were in close contact with infected individuals and may be at risk of infection. We assumed that a proportion of individuals would be voluntarily tested according to the coverage rate (denoted as $\tau$). Susceptible individuals (S) who have undergone voluntary testing would return to the susceptible compartment (S) after the testing to diagnosis interval. In contrast, asymptomatic/pre-symptomatic infected individuals who have undergone voluntary testing would be diagnosed after the testing to diagnosis interval and thus strictly isolated.

Vaccination would reduce the proportion of susceptible individuals in the population. Individuals who have been vaccinated and have developed immune protection would enter the vaccination compartment (V). We assumed that this population would not be able to be infected in the short term.

#### 2. Data and parameter estimation

##### 2.1 Epidemiological data

We searched historical outbreak data for COVID-19 from the official website of the Australian Department of Health for the period 25 January 2020 to 12 March 2021, including the number of daily reported cases (both with known and unknown sources), cumulative confirmed cases, and deaths. Because some states and territories did not report information on the source of confirmed cases (e.g., whether cases were from known clusters), satisfactory data from Victoria, New South Wales, the Australian Capital Territory, and Western Australia were collected for analysis. We calibrated the model with relevant data from Victoria. Further, we verified the reliability of the model outputs with relevant data from New South Wales, the Australian Capital Territory, and Western Australia, respectively.

The epidemiological data from the four states mentioned above were presented in Supplementary Fig. 2. Confirmed cases are classified according to their source as overseas cases, locally known cases and locally unknown cases. From 16 March 2020, all arrivals in Australia were required to be in self-imposed isolation for 14 days, which became mandatory from 28 March 2020. Therefore, we considered that the activity of overseas cases was unrestricted until 16 March 2020, and overseas cases had the same potential for community transmission as locally acquired cases of known sources. We assumed that 50% of overseas cases might comply with strict isolation between 17 March 2020 and 28 March 2020. We also assumed that all overseas cases might be in strict isolation after 29 March 2020, so they contributed only to the number of cases but not community transmission.

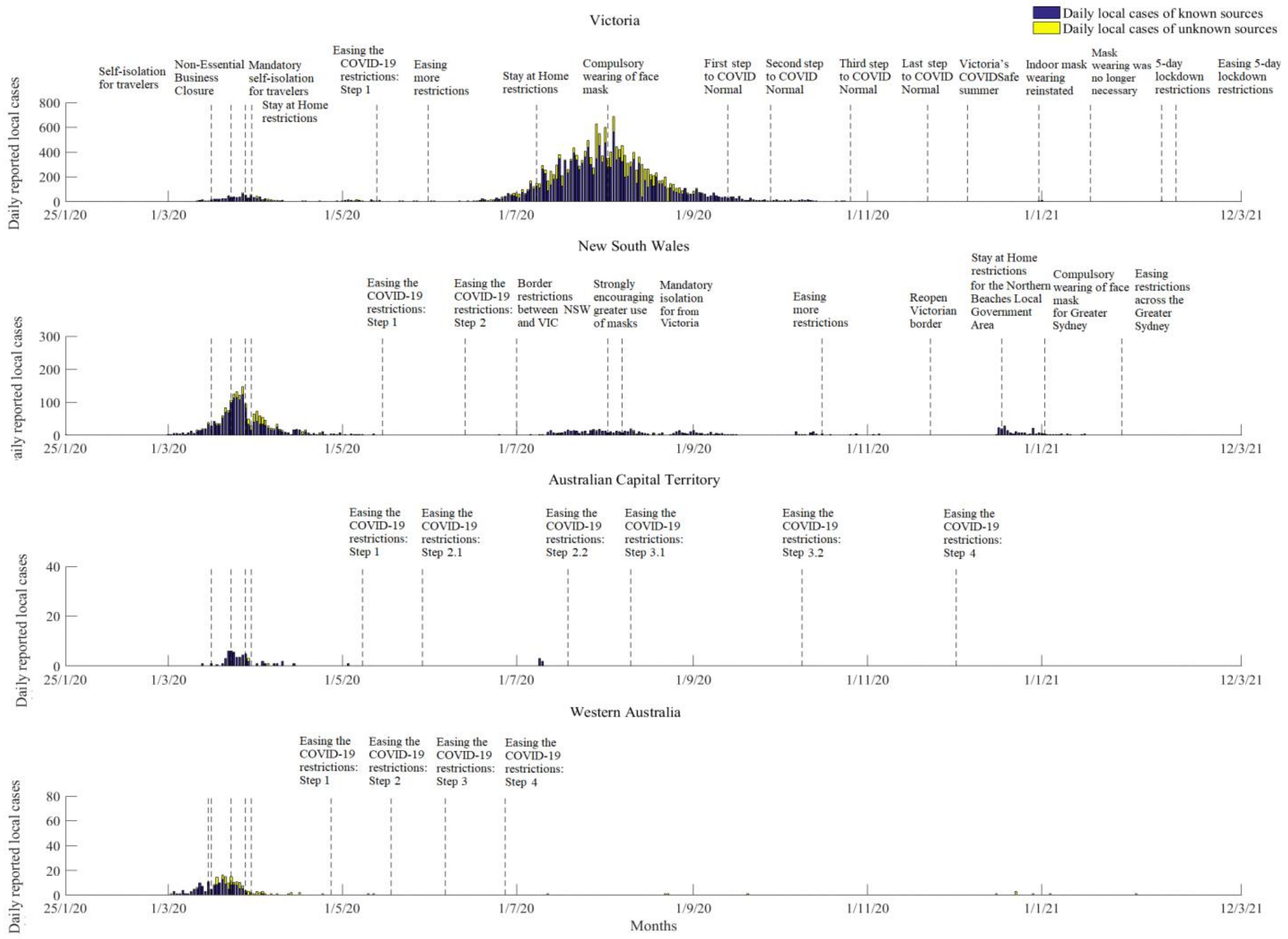


**Supplementary Fig. 2. COVID-19 epidemiological data and public health interventions in four Australian states (25 January 2020 – 12 March 2021)**

Supplementary Fig. 3 illustrated the proportion of daily unknown-source cases to total daily locally acquired cases in the historical outbreaks in the four states. We found that about 20% of locally acquired cases in Australia were of unknown sources. This proportion had large fluctuations, especially when the number of daily cases was low.

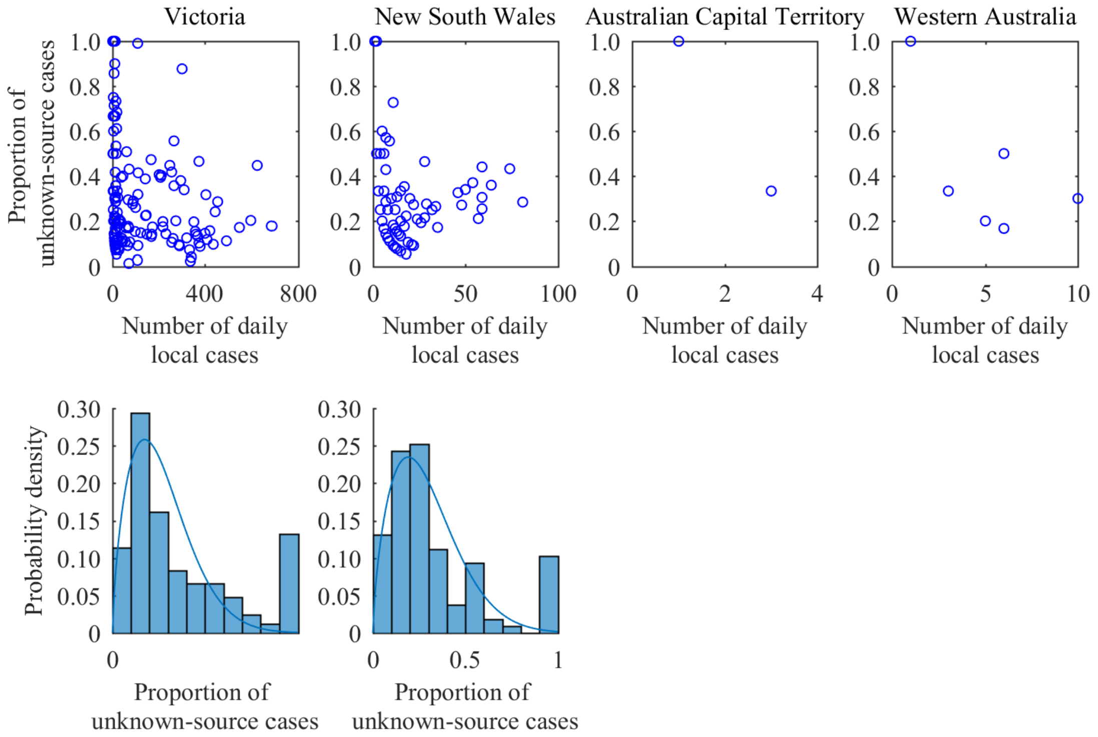


**Supplementary Fig. 3 The proportion of unknown-source cases to total locally acquired cases in historical outbreaks in four Australian states**

#### 2.2 Data related to public health interventions

##### (1) Reduction in social activity and average number of daily close contacts

The core elements and timelines of relevant public health policies implemented by each Australian state to control the spread of SARS-CoV-2 were collected and presented in Supplementary Fig. 2. To assess the impact of social distancing restrictions on social activities, we analyzed Google COVID-19 community mobility data and obtained changes in mobility in public places[4]. We expressed mobility changes as proportional deviations from levels for the baseline (Supplementary Fig. 4). It can be seen that policies influenced mobility in public places and that mobility decreased as restrictions were imposed and increased as restrictions were relaxed.

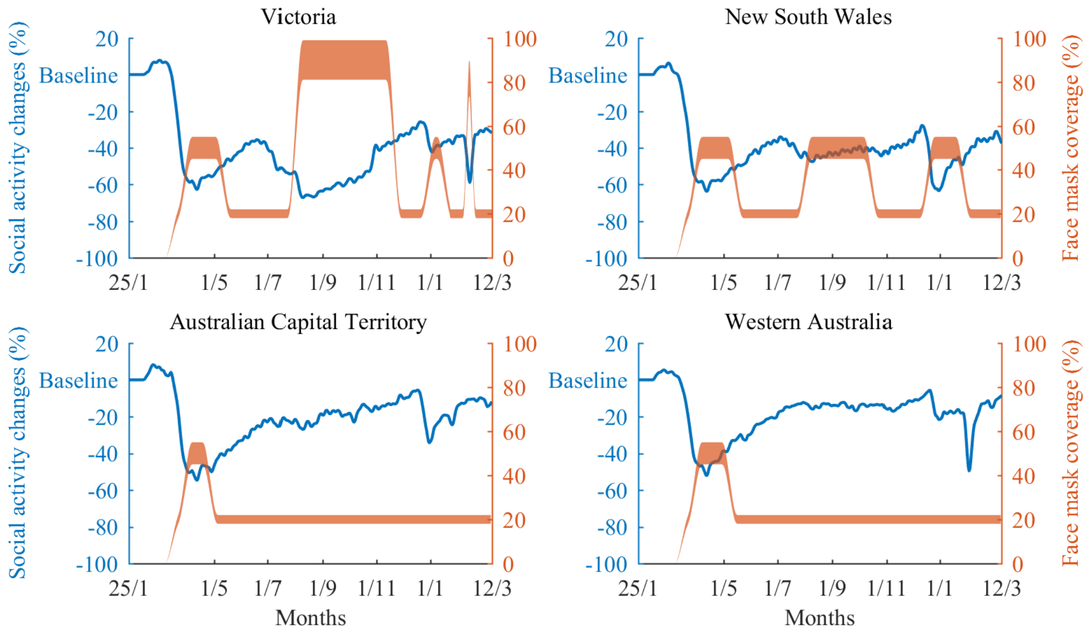


**Supplementary Fig. 4 Social activity changes and face mask coverage rates in four Australian states (25 January 2020 – 12 March 2021)**

We estimated that the average daily number of close contacts for individuals in public places in Australia without social distancing restrictions was 19, based on previous reports[5]. Further, we estimated the real-time average daily number of close contacts in public places based on the mobility changes in public places to simulate the impact of social distancing restrictions. According to the Australian Bureau of Statistics, the average household size is three. Therefore, the average number of close contacts in a household was estimated to be two.

##### (2) Effectiveness and coverage rate of face mask use

The effectiveness of face mask use in preventing infection was estimated to be 85% (95% CI: 50–95%), based on relevant meta-analysis against COVID-19[6,7]. In the context of the COVID-19 pandemic, the rate of face mask use in public places in Australia is around 10–30%, according to the report from the global health research centre at the University of Washington[8]. We assumed that face mask use in public places would increase spontaneously to about 50% when the lockdown was implemented. During mandatory face mask measures in Victoria, the face mask coverage in public places was estimated to reach 80–100%. The ranges of face mask coverage rates over time were estimated based on the relevant public health policies and were displayed in Supplementary Fig. 4.

##### (3) Effectiveness of contact tracing, rate of voluntary testing and vaccine efficacy

The effectiveness of contact tracing may depend on various factors, such as willingness to cooperate, recall bias, availability of contact tracers, and quarantine compliance. We estimated that 20% of the close contacts would be uncooperative, and among those who cooperate, 60% of recollected information may be incorrect[2,3]. A further 20% of close contacts would likely fail to comply with quarantine[2]. We estimated that approximately 80% of close contacts among the remaining close contacts (cooperative and without recall bias) would be detected and isolated in Australia using model calibration.

Based on the cumulative number of COVID-19 voluntary tests over the past 7 days and the population size reported by the Australian Government Department of Health, we estimated that 0.09%–0.2% of the Australian population would receive voluntary testing each day.

According to Australia's vaccine agreements, the Pfizer/BioNTech vaccine (40 million doses available), the Oxford/AstraZeneca vaccine (53.8 million doses available), and the Moderna vaccine (25 million doses available) will account for 33.7%, 45.3%, and 21.0% of COVID-19 vaccination in Australia, respectively[9]. The efficacy of the Pfizer/BioNTech vaccine, the Oxford/AstraZeneca vaccine, and the Moderna vaccine has been reported to be 95% (90.3–97.6%), 67.1% (52.3–77.3%), and 94.1% (89.3–96.8%), respectively[11–12]. We hence estimated the weighted population vaccination effectiveness to be about 82.2%. Recent studies have shown that the existing vaccines remain equally effective in preventing clinical severities in patients infected with Alpha and Delta variants. Still, there was a slight decrease in effectiveness against infection. The efficacy of the Pfizer/BioNTech vaccine reduced to 93.4% (90.4–95.5%) for Alpha and 87.9% (78.2–93.2%) for Delta variant[13–15]. The efficacy of the Oxford/AstraZeneca vaccine reduced to 66.1% (54.0–75.0%) for Alpha and 59.8% (28.9–77.3%) for Delta variant[13–15]. We, therefore, assumed a 2% and 10% reduction in the efficacy of the vaccine against Alpha and Delta.

##### 2.3 Transition probability between model compartments

Transition probabilities in the model were derived from published literature and model calibration and were summarized in Supplementary Table 1. We first obtained plausible initial ranges for uncertain parameters through an extensive literature review and then obtained the good-fitting parameter sets by model calibration. Multiple calibration targets were established for model calibration, including the number of daily confirmed cases, the number of daily known-source cases, the number of daily unknown-source cases, and cumulative deaths, from historical outbreaks in Victoria. We calibrated the model by comparing the model predictions with these calibration targets and deducing the most probable values of the set of parameters. In this process, we used a calibration procedure using the genetic algorithm[16]. We performed about 1,000,000 simulations in total. In each simulation, one value for each parameter was randomly extracted from its initial range, and a set of input values was formed. With this set of input values, the compartmental model was run to produce the outputs compared with the calibration targets. A goodness-of-fit score was calculated by summing the log-likelihoods. Goodness-of-fit scores were assumed to follow a chi-square distribution with the number of degrees of freedom equal to the number of calibration targets. Based on this distribution, 'good-fitting sets' of model parameters were identified using the likelihood ratio test, comprising those sets that did not produce a non-inferior fit compared with the best-fitting set (using an alpha level of 5%)[17]. We ranked the goodness-of-fit scores of the 'good-fitting sets' in ascending order and retained the top 1000 sets of best goodness-of-fit. The best-fitting set was the set with the lowest goodness-of-fit score whose simulated outputs were closest to the calibration targets (Supplementary Table 1). The best-fitting set was introduced into the model as the base-case values of parameters, and the 1000 'good-fitting sets' were used for sensitivity analyses. All analyses and simulations were performed in MATLAB R 2019a.

**Supplementary Table 1 Transition probabilities between model compartments**

| Parameter | Initial range and reference | Best-fitting set |
|---|---|---|
| The average probability of being infected per day by contact with a symptomatic infected individual in the household ($\beta$) | 0.01 – 0.05[18] | 0.0285 |
| Percentage reduction in the average daily probability of being infected by contact with an infectious individual in a public place compared to that of being infected by contact with an infectious individual in households ($\rho$) | 0 – 1 | 0.60 |
| The reduction in daily transmission probability by contact with an asymptomatic/pre-symptomatic infected individuals ($\varepsilon$) | 0 – 1 | 0.275 |
| The proportion of asymptomatic infections among newly infected individuals ($\psi$) | 0.101 – 0.23[19,20] | 0.174 |
| The mean incubation time (days) ($1/v$) | 5.0 – 6.7[21] | 5.8 |
| The interval from symptom onset to isolation in hospital or quarantine (days) ($1/q$) | 2 – 8[22] | 4.2 |
| The interval from testing to diagnosis (days) ($1/g$) | 1 – 3 | 2.3 |
| The mean time from infection to recovery for asymptomatic infected individuals (days) ($1/r_1$) | 11 – 26[23] | 22 |
| The mean time from diagnosis to recovery for symptomatic infected individuals (days) ($1/r_2$) | 11 – 26[23] | 17 |
| Average daily probability of death due to disease during treatment ($\mu$) | 0.001 – 0.005[18] | 0.002 |

For illustrative purposes, we simulated the historical epidemic trends in Victoria based on the state's total population using the 1000 'good-fitting sets'. We yielded the number of daily confirmed cases, the number of daily unknown-source cases, and the cumulative number of deaths, respectively. The outputs were compared with the corresponding calibration targets, as shown in Supplementary Fig. 5.

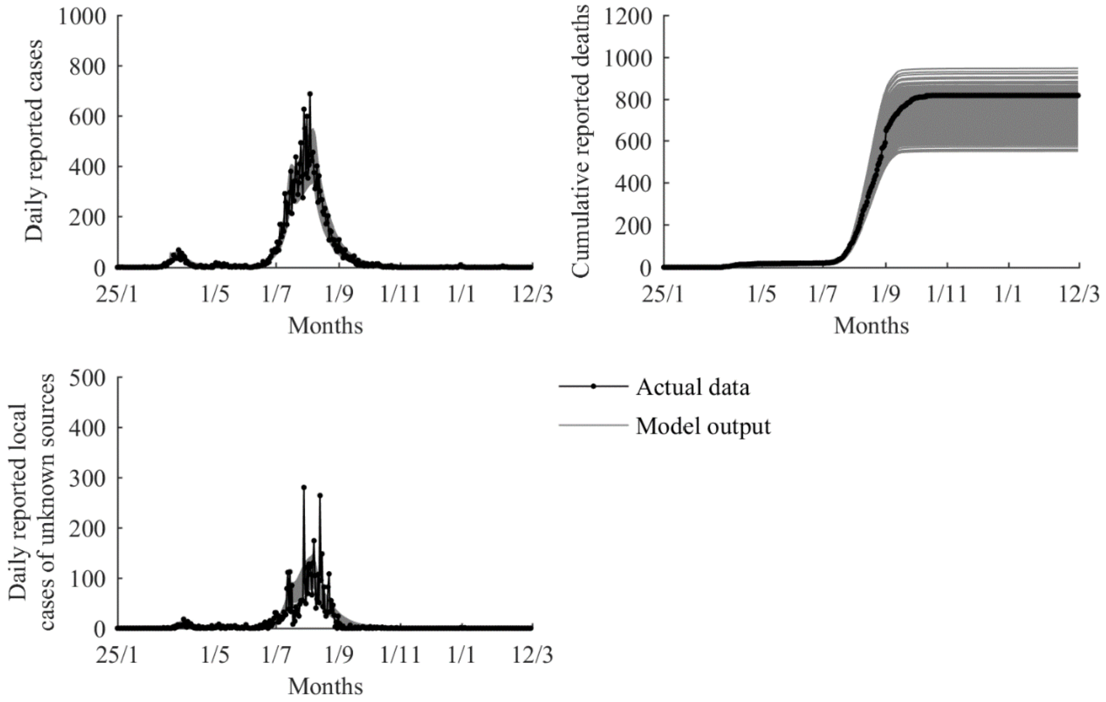


**Supplementary Fig. 5 Model outputs of COVID-19 outbreak trends in Victoria (25 January 2020 – 12 March 2021)**

### 3. Effective reproduction number

The effective reproduction number ($R_e$), the average number of secondary infections caused by a single infective at a given susceptible fraction, is calculated as the largest eigenvalue of the next generation matrix K = F×V$^{-1}$ [24–26], where

$F=$

$$\begin{bmatrix} \left(\beta_{EA}^{fam}\cdot c_f+\beta_{EA}^{pub}\cdot c_p(t)\right)\cdot\psi & \left(\beta_{EA}^{fam}\cdot c_f+\beta_{EA}^{pub}\cdot c_p(t)\right)\cdot\psi & \left(\beta_{I}^{fam}\cdot c_f+\beta_{I}^{pub}\cdot c_p(t)\right)\cdot\psi \\ \left(\beta_{EA}^{fam}\cdot c_f+\beta_{EA}^{pub}\cdot c_p(t)\right)\cdot(1-\psi) & \left(\beta_{EA}^{fam}\cdot c_f+\beta_{EA}^{pub}\cdot c_p(t)\right)\cdot(1-\psi) & \left(\beta_{I}^{fam}\cdot c_f+\beta_{I}^{pub}\cdot c_p(t)\right)\cdot(1-\psi) \\ 0 & 0 & 0 \end{bmatrix}$$

and

$V=$

$$\begin{bmatrix} (r_1+\tau)+\begin{pmatrix}\beta_{EA}^{fam}\cdot\xi\cdot g\cdot(c_f-1)+\\ \beta_{EA}^{pub}\cdot\xi\cdot g\cdot c_p(t)\end{pmatrix}\cdot\psi\cdot\xi & \begin{pmatrix}\beta_{EA}^{fam}\cdot\xi\cdot g\cdot(c_f-1)+\\ \beta_{EA}^{pub}\cdot\xi\cdot g\cdot c_p(t)\end{pmatrix}\cdot\psi\cdot\xi & \begin{pmatrix}\beta_{I}^{fam}\cdot q\cdot(c_f-1)+\\ \beta_{I}^{pub}\cdot q\cdot c_p(t)\end{pmatrix}\cdot\psi\cdot\xi \\ \begin{pmatrix}\beta_{EA}^{fam}\cdot\xi\cdot g\cdot(c_f-1)+\\ \beta_{EA}^{pub}\cdot\xi\cdot g\cdot c_p(t)\end{pmatrix}\cdot(1-\psi)\cdot\xi & (v+\tau)+\begin{pmatrix}\beta_{EA}^{fam}\cdot\xi\cdot g\cdot(c_f-1)+\\ \beta_{EA}^{pub}\cdot\xi\cdot g\cdot c_p(t)\end{pmatrix}\cdot(1-\psi)\cdot\xi & \begin{pmatrix}\beta_{I}^{fam}\cdot q\cdot(c_f-1)+\\ \beta_{I}^{pub}\cdot q\cdot c_p(t)\end{pmatrix}\cdot(1-\psi)\cdot\xi \\ 0 & -v & q \end{bmatrix}$$

The effective reproduction number under vaccination $R_v$ is the number of secondary cases caused by one primary case introduced into a certain proportion of the vaccinated population[27,28]. We assumed that individuals who receive the vaccine and develop an immune response would no longer be infected over a period of time. Thus, $R_v$ was expressed as:

$$R_V=(1-\theta\cdot p)\cdot R_e \quad (7)$$

Where $\theta$ denotes the efficacy of the vaccine and $p$ denotes the vaccination coverage.

### 4. Estimation of undocumented cases

Undocumented cases represented a potential risk of further community transmission of SARS-CoV-2. Three types of infections were considered 'undocumented' in our model. They were asymptomatic infections, pre-symptomatic infections, and symptomatic infections before diagnosis. The number of undocumented cases in historical outbreaks in Victoria was estimated based on the compartmental model and was shown in Supplementary Fig. 6. As can be seen, the cumulative number of cases rose at the fastest rate when the number of active undocumented cases reached a peak.

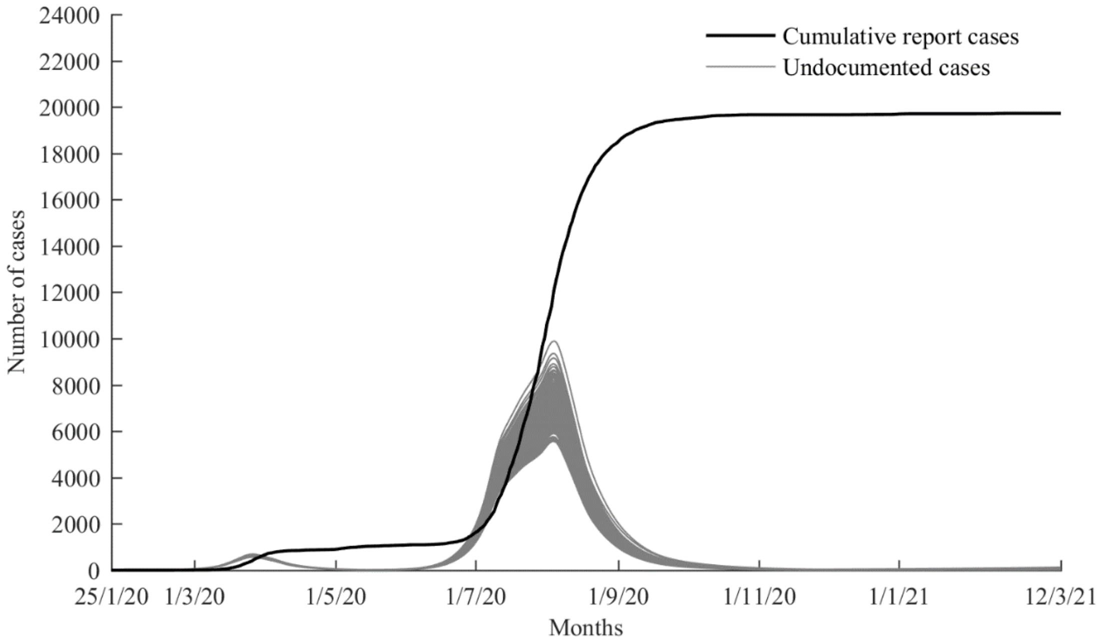


**Supplementary Fig. 6 Model outputs of undocumented cases (including asymptomatic infected individuals, pre-symptomatic infected individuals, and symptomatic infected individuals before diagnosis) in historical outbreaks in Victoria (25 January 2020 – 12 March 2021)**

### 5. Association between the number of undocumented cases and the number of daily reported cases

Supplementary Fig. 7 demonstrated the potential reasons for cases being identified as unknown sources. Supplementary Fig. 8 showed the relationship between the number of daily unknown-source cases and the number of active asymptomatic infections, and the relationship between the number of daily known-source cases and the number of active pre-symptomatic/symptomatic infections. The number of active undocumented cases was estimated from the model, and the number of daily cases was obtained from reported data. Using the 1000 'good-fitting sets' of model parameters, we calculated the interquartile range of the slope of the linear relationships, which was shown in Supplementary Fig. 9.

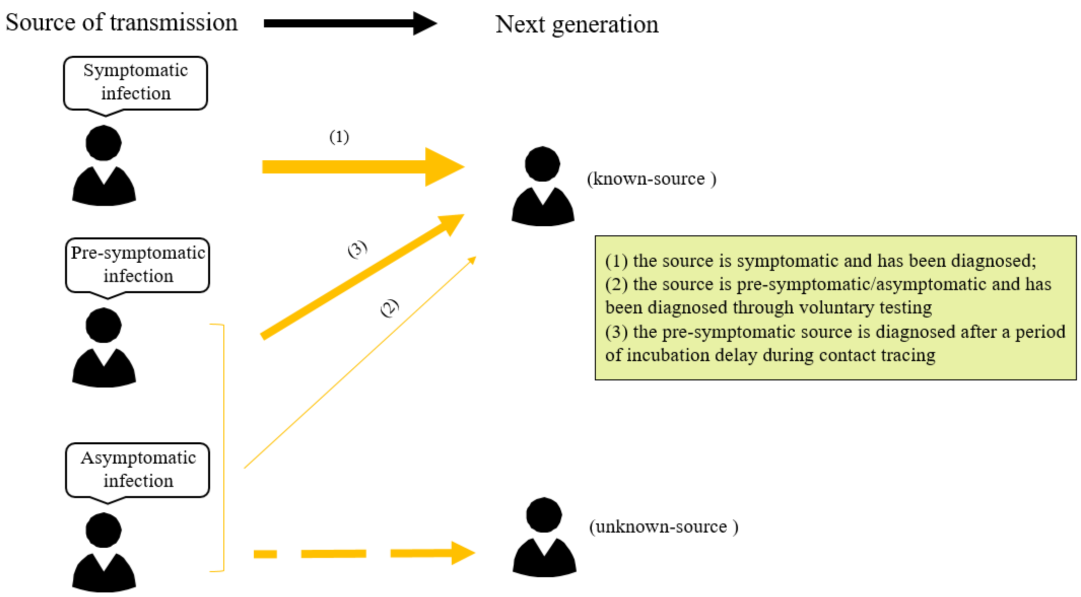


**Supplementary Fig. 7 The reasoning process for the source of confirmed cases**

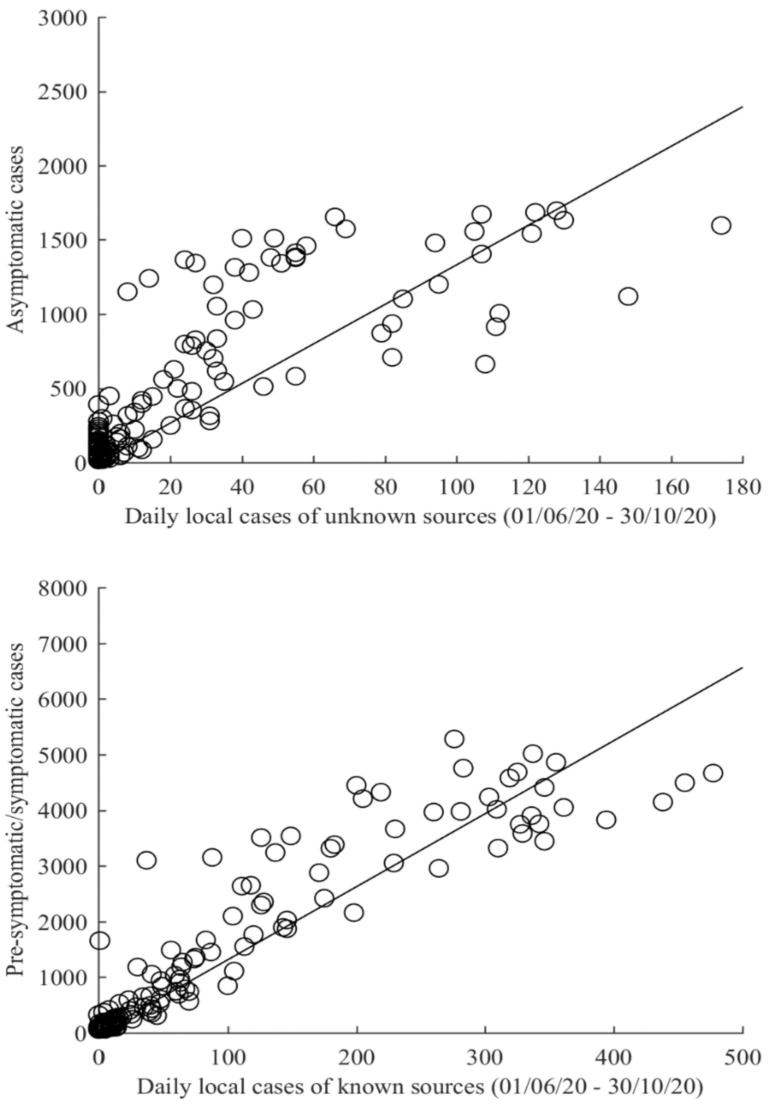

**Supplementary Fig. 8 The linear relationships between the number of daily unknown-source cases and the number of model-estimated asymptomatic infections, and between the number of daily known-source cases and the number of model-estimated pre-symptomatic/symptomatic infections**

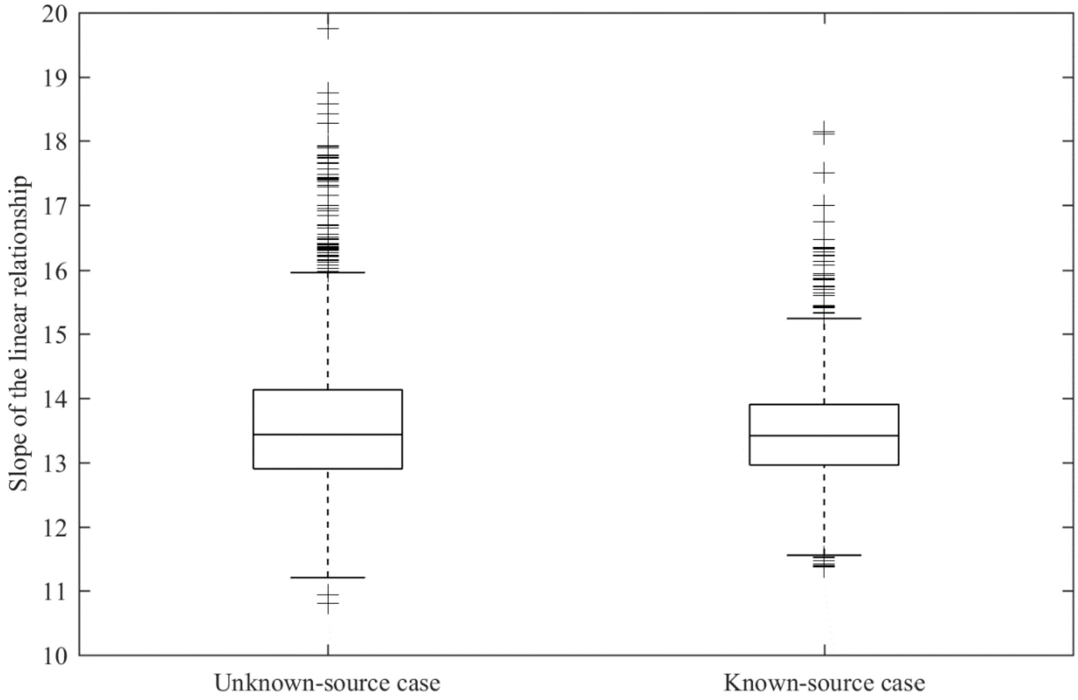


**Supplementary Fig. 9 Sensitivity analysis for the linear relationships**

### 6. Predicted number of reported cases over the next 7 days

Based on the definition of $R_e$, i.e., the number of secondary cases generated by a single infectious case, we multiplied $R_e$ with the number of undocumented cases to obtain the total number of secondary cases caused by the current source of infection during the average infectious period.

We estimated the average infectious period for individuals infected with SARS-COV-2 as follows. Previous studies have indicated that the mean incubation period for pre-symptomatic infected individuals (who later became symptomatic) was approximately 5.8 (95% CI 5.0–6.7) days[21], and the interval from symptom onset to isolation in hospital or quarantine was about 5.6 (IQR 2–8) days[22]. Therefore, the average infectious period for a pre-symptomatic case was approximately 11.4 days. In contrast, asymptomatic infected individuals who never presented any symptoms would additionally experience an asymptomatic recovery period of $17 \pm 4$ (range 11–26) days[23], resulting in an overall infectious period (i.e. incubation period plus asymptomatic recovery period) of 22.8 days. Given the proportion of asymptomatic cases among infected cases is 17% (95% CI 14–20%)[19], the weighted average infectious period of a SARS-COV-2 infected individual was estimated to be about 14 days.

According to the linear relationships (Supplementary Fig. 8–9), we estimated the number of undocumented cases in the populations based on the number of daily reported cases. Then, the overall number of secondary cases over the next 14 days was obtained by multiplying $R_e$ with the number of undocumented cases. We divided this number by 2 to obtain the number of reported cases over the next 7 days.

#### 7. Model validation

Using the method described above, we predicted the number of reported cases in the next 7 days based on the average number of daily cases in the first three days of historical epidemic data and formed prediction curves for epidemic trends in Victoria, New South Wales, the Australian Capital Territory, and Western Australia, respectively. Supplementary Fig. 10 showed the comparison between the projected number of cases over the next 7 days with the actual number of reported cases over a 7-day period in each of Victoria, New South Wales, Australian Capital Territory, and Western Australia, for the period 25 January 2020 –12 March 2021. It can be seen that the model predictions matched well with the actual epidemic trends, with an R-squared of 0.99, 0.85, 0.74, and 0.87, respectively.

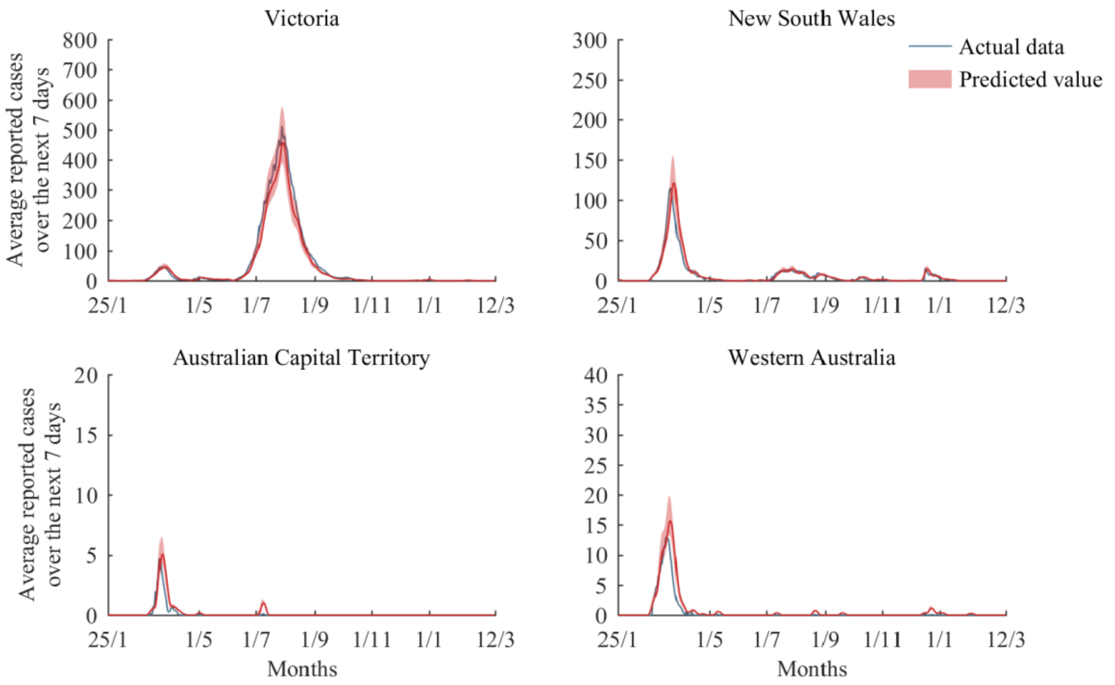


**Supplementary Fig. 10 Model predictions of COVID-19 outbreak trends in Victoria, New South Wales, the Australian Capital Territory, and Western Australia (25 January 2020 –12 March 2021)**
